# Aids to Improve Statistical Risk Communication in Patients Consenting for Surgery and Interventional Procedures: a Systematic Review

**DOI:** 10.1101/2023.01.19.23284521

**Authors:** Arif HB Jalal, Despoina Chatzopoulou, Hani J Marcus, Anand S Pandit

## Abstract

**Objective:** Evaluate the effect of risk communication tools on the understanding of statistical risk of complications occurring in patients undergoing a surgical or interventional procedure.

**Summary Background Data:** Informed consent is an essential process in clinical decision-making, through which healthcare providers educate patients about the benefits, risks and alternatives of a procedure. Numerical risk information is by nature probabilistic and difficult to communicate. Aids which support statistical risk communication and studies assessing their effectiveness are needed.

**Methods:** A systematic search was performed across Medline, Embase, PsycINFO, Scopus and Web of Science until July 2021 with a repeated search in September 2022. Studies examining risk communication tools (e.g. informative leaflets, audio-video) in adults (age>16) patients undergoing a surgical or interventional procedure were included. Studies only assessing understanding of non-statistical aspects of the procedure were excluded. Both randomised control trials (RCTs) and observational studies were included. Cochrane risk-of-bias and the Newcastle-Ottawa Scale were used to assess the quality of studies. Due to heterogeneity of the studies, a narrative synthesis was performed (PROSPERO ID: CRD42022285789).

**Results:** A total of 4348 articles were identified and following abstract and full-text screening a total of 11 articles were included. 8 studies were RCTs and 3 were cross-sectional. The total number of adult patients was 1030. The most common risk communication tool used was additional written information (n=7). Of the 8 RCTs, 5 showed statistically significant improvements in the intervention group in outcomes relating to recall of statistical risk. Quality assessment of RCTs found some concerns with all studies.

**Conclusions:** Risk communication tools appear to improve recall of statistical risk. Additional prospective trials are warranted which can compare various aids and determine the most effective method of improving patient understanding.

## INTRODUCTION

Informed consent represents an essential process in clinical decision-making, through which healthcare providers educate patients about the benefits, risks and alternatives of a given procedure or intervention in a descriptive way. Statistical risk information pertaining to medical procedures is by nature probabilistic and can be difficult to communicate. For cognitively intact patients, healthcare professionals typically narrate the chance of a complication occurring either in descriptive terms such as ‘common’, ‘uncommon’ or ‘rare’ or by means of a percentage. Occasionally proportional descriptors are used to relay mathematical information such as *“9 in 10 patients will proceed without a complication”* rather than saying there is a *“10% risk of suffering one”*. However, narrating the statistical chances of a complication occurring in these ways may be fraught with pitfalls, at least in part due to the limited understanding of health and lack of numerical literacy among patients, including those undergoing surgery. Indeed, a recent metanalysis of 18,895 surgical patients from 40 studies showed a pooled estimate prevalence of 31.7% for limited health literacy^1^. Numeracy, defined as “the ability to understand and use numbers in daily life”^2-4^, in particular, is often poor among the general population, with the majority of adults having difficulty converting small frequencies such as ‘1 in 1000’ to 0.1% ^5^.

Ordinarily during the surgical consent process, the numerical skills of patients are not evaluated nor is the presentation of medical information adapted to the patients’ educational level. There is also considerable variation regarding how numerical probabilities (e.g. negligible, low) are translated into verbal probabilities among clinicians^6^. In addition, the recall of surgical complications following consent is poor ^7-9^. Patients undergoing spinal or cranial surgery, for example, are able to retain just 18% of the information regarding operative risks, when assessed just 2 hours after consent is obtained ^7^. Furthermore, numerical risks are often presented in ambiguous terms, and it is unclear whether quoted statistics are cumulative (with reference to all the possible complications) rather than for individual events. Finally, even for those with adequate statistical understanding, relating objective probabilities in a personal way is difficult and often requires additional heuristics to facilitate greater understanding^10^.

Decision aids describe a “means of helping people make informed choices about healthcare that take into account their personal values and preferences” ^11^ and may help in the communication of numerical risks (**Figure 1)**. While they have previously been used in settings related to screening, clinical trials or communicating risk of disease^12-14^, it is unclear the extent to which they are used for patients consenting for surgery. Surgery for many is a life event that carries a significant emotional burden at a time of suffering and debilitation, and aids which can help navigate the intended treatment therefore have clear potential. Patient information sheets, structured interview techniques, videos/multimedia and visual aids have been previously studied in relation to the assessment of understanding of surgical procedures ^15-19^. However, many of these studies measure the ability of patients to recall the names of specific complications rather than their associated numerical or probabilistic risk.

**Figure 1.**
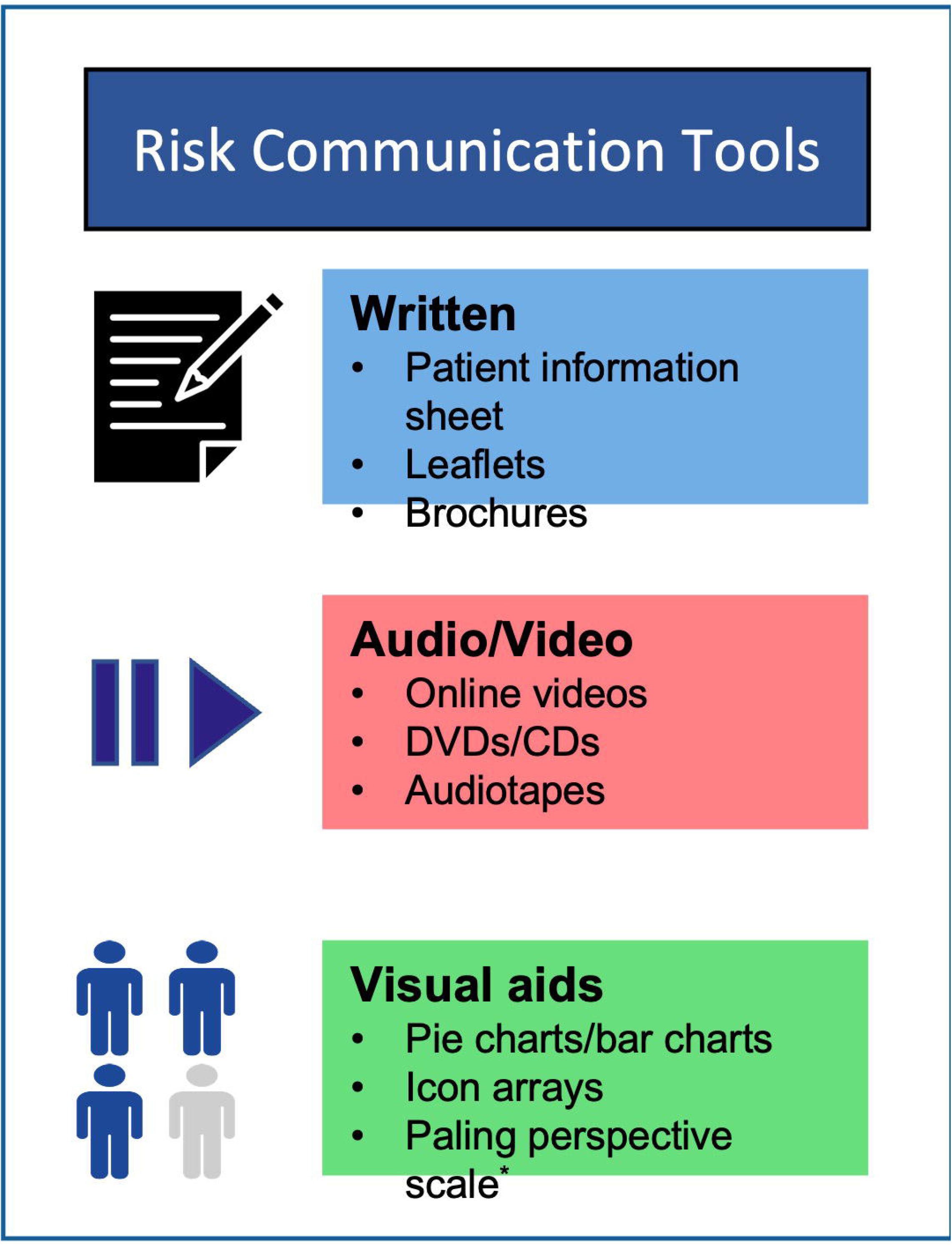
Examples of different risk communication tools. Figure designed using Flaticon.com with icons from Iakonicon and Dinosoft labs. * Paling perspective scale is a graphical method used to communicate risks.^59^

While the effectiveness of risk communication methods has been compared in medical populations ^14,20^, no such review, to the best or our knowledge, focuses on patients undergoing surgery. To that end, this systematic review aims to study the impact of aids and other risk communication adjuncts on numerical understanding and perception of probabilistic risk in patients undergoing surgery and consent-requiring interventional procedures.

## METHODS

This systematic review was designed in accordance with the Preferred Reporting Items for Systematic Reviews and Meta-analyses (PRISMA) reporting guideline and registered on PROSPERO (CRD42022285789)^21^.

### Search strategy

#### Eligibility Criteria

Eligibility criteria of studies was developed using the population, intervention, comparator and outcome (PICO) framework. The population of interest was any adult population undergoing an invasive surgical or consent-requiring interventional procedure. The latter being defined as procedures used for diagnosis or treatment that involve incision, puncture, entry into a body cavity or the use of ionising, electromagnetic or acoustic energy ^22^. For the purpose of this study, the ‘intervention’ of interest was defined as exposure of patients to a decision aid, tool, method or consent adjunct aimed at improving a patient’s understanding of statistical information relating to the procedure. Relevant outcomes would be any assessment of the recall, understanding, sentiment or perception of probabilistic or numerical risks related to a procedural complication. Observational studies discussing the patients’ knowledge of statistical risk were also included. Studies involving aids or processes solely aimed at helping the patient understand non-statistical aspects of the procedure were excluded.

#### Information Sources

Search terms were refined and then a systematic search was performed by the study team and an academic librarian with over 20 years’ experience on July 13^th^ 2022. In total, five databases were included: Ovid Medline, Embase, APA PsychInfo, Scopus and Web of Science. Results were de-duplicated and exported to EndNote X9.3.3 (Philadelphia, PA, USA)^23^. Additional articles were identified through the reference list of relevant reviews. Both English and non-English articles were included. Details of the search strategy used can be found in *supplementary 1*. The search was repeated on 5^th^ September 2022.

#### Study Selection

Abstracts of all articles were independently screened by two reviewers (AHBJ and DC). Disagreements were resolved through discussion and with a third reviewer (ASP). Full-text screening was performed in the same manner.

#### Data Extraction and Synthesis

##### Data Extraction

Data from all included articles were independently extracted by two individuals (AHBJ and DC) using a pre-defined data collection form. Data extracted included study author, study design, sample size, age range, population studied, interventions used, comparator groups used, measured outcomes, method of assessing probabilistic understanding and results relating to our review question.

##### Quality Assessment

Randomised control trials were assessed using the Cochrane Risk-of-Bias Tool^24^ while cross-sectional studies were assessed using a version of the Newcastle-Ottawa Scale that had previously been adapted for cross-sectional studies.^25^ Two reviewers (AHBJ and DC) each independently assessed the quality of the included studies using the relevant criteria. Any disagreements were resolved through discussion. Certainty of evidence was assessed using the GRADE approach ^26^.

##### Data Synthesis

A narrative synthesis was performed in accordance to the Synthesis Without Meta-Analysis guideline.^27^ This was chosen in favour of a meta-analysis due to heterogenous outcome reporting amongst studies. Studies were grouped based on the type of intervention used (e.g. videos, written information, visual aids). Summaries of effects for comparative studies were intended to be reported using risk ratios (for binary outcomes) and standardised mean differences (for continuous outcomes) where possible

## RESULTS

### Identification of Eligible Studies

A PRISMA flow diagram summarizing our search results is presented in **Figure 2**. Our initial search identified 724 records from Medline, 3183 records from Embase, 167 records from PsycInfo, 1231 records from Scopus and 824 records from Web of Science. Following de-duplication and screening, 11 studies were included for data extraction.

**Figure 2.**
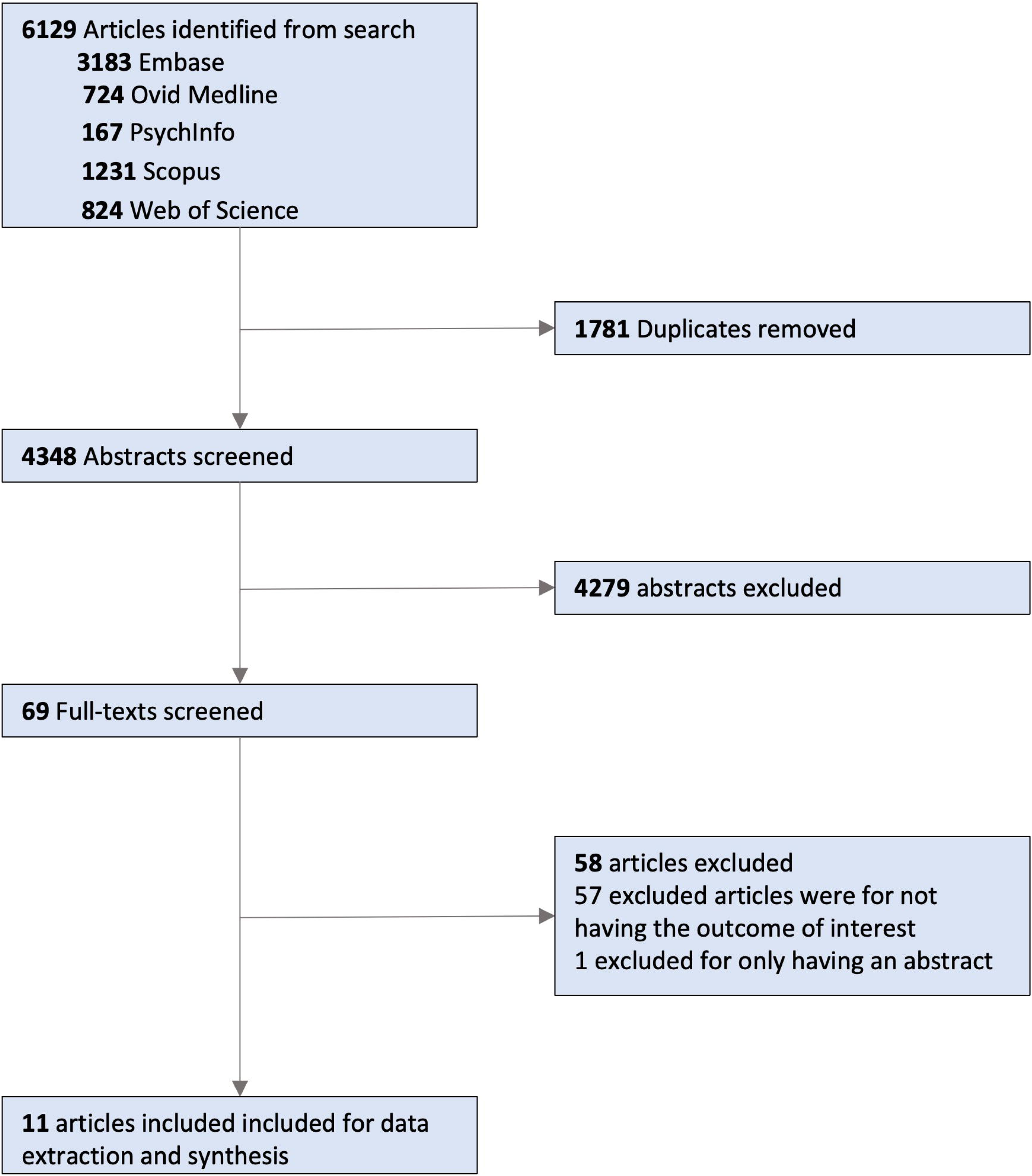
PRISMA flow diagram

### Characteristics of Studies

The characteristics and outcomes of included studies are summarized in **Table 1** and **Table 2**. In total, 11 studies, with a total sample size of 1051 participants, were identified which made attempts at objective assessment of numerical or probabilistic understanding of risk in patients undergoing surgery or interventional procedures. As one study included children in its sample size the total number of adults is 1030 ^28^. Most of the studies were randomised control trials (n = 8, 72.7%) with the remainder being cross-sectional studies (n = 3, 27.3%).

**Table 1.** Study design characteristics of included studies. RCT = Randomised controlled trial, CABG = Coronary artery bypass graft, ERCP = Endoscopic retrograde cholangiopancreatography

| Study | Study Design | Sample size (control) | Mean (Age range) | Procedure | Decision Aid Medium | Intervention | Control |
| --- | --- | --- | --- | --- | --- | --- | --- |
| Alsaffar et al.<sup>29</sup> | RCT | 49 (25) | 49.0 (27-77) | Total thyroidectomy | Written | Written information sheet + verbal consent | Verbal consent |
| Bhambhwani et al.<sup>30</sup> | RCT | 28 (14), 21 children | 45.8 (N/A), adults | Strabismus surgery | Written | Written information sheet + verbal consent | Verbal consent |
| Gett et al.<sup>31</sup> | Cross-sectional | 32 | 48.3 (21-73) | Colonoscopy | Graphical | Graphical formats: Pie chart, 1000-person diagram (icon array), logarithmic scale<br>Text formats: Absolute risk ratio, relative risk ratio | N/A |
| Habib et al.<sup>32</sup> | RCT | 200 (100) | 68.8 (37-94) | Peripheral angioplasty | Written & Graphical | Risk assessment chart (patient information sheet provided based on referral pathway) | No chart (patient information sheet provided based on referral pathway) |
| Inglis et al.<sup>33</sup> | RCT | 40 (20) | 46.5 (21-80) | Surgery requiring general anaesthesia | Written & audio/video | Audiotape recording + written information sheet (with detailed information) | Audiotape recording + written information sheet (with routine information) |
| Laupacis et al.<sup>34</sup> | RCT | 120 (61) | 60.0 (20-81) | Elective cardiovascular surgery (CABG, valve surgery or combined) | Written & audio/video | Audiotape + booklet. | Routine information |
| Lee et al.<sup>35</sup> | Cross-sectional | 126 | 53.2 (N/A) | Mastectomy | Unspecified | Verbal consent | N/A |
| Lloyd et al.<sup>36</sup> | Cross-sectional | 71 | Unspecified | Carotid endarterectomy | Unspecified | Unspecified | N/A |
| Shukla et al.<sup>37</sup> | RCT | 100 (25) | 74 (N/A) | Cataract surgery | Written & audio/video | Group 2: verbal information + 2<sup>nd</sup> grade reading level written information sheet;<br>Group 3: verbal information + 8<sup>th</sup> grade reading level brochure;<br>Group 4: verbal information + patient education video | Verbal consent (Group 1) |
| Winfield et al.<sup>38</sup> | RCT | 80 (38) | N/A (19-68) | Excretory urography | Written | Detailed written informed consent form + interview | Verbal consent |
| Xia et al.<sup>39</sup> | RCT | 205 (104) | 43.3 (N/A) | ERCP | Audio/video | Patient education video detailing procedure + standard written informed consent | Written informed consent |

**Table 2.**
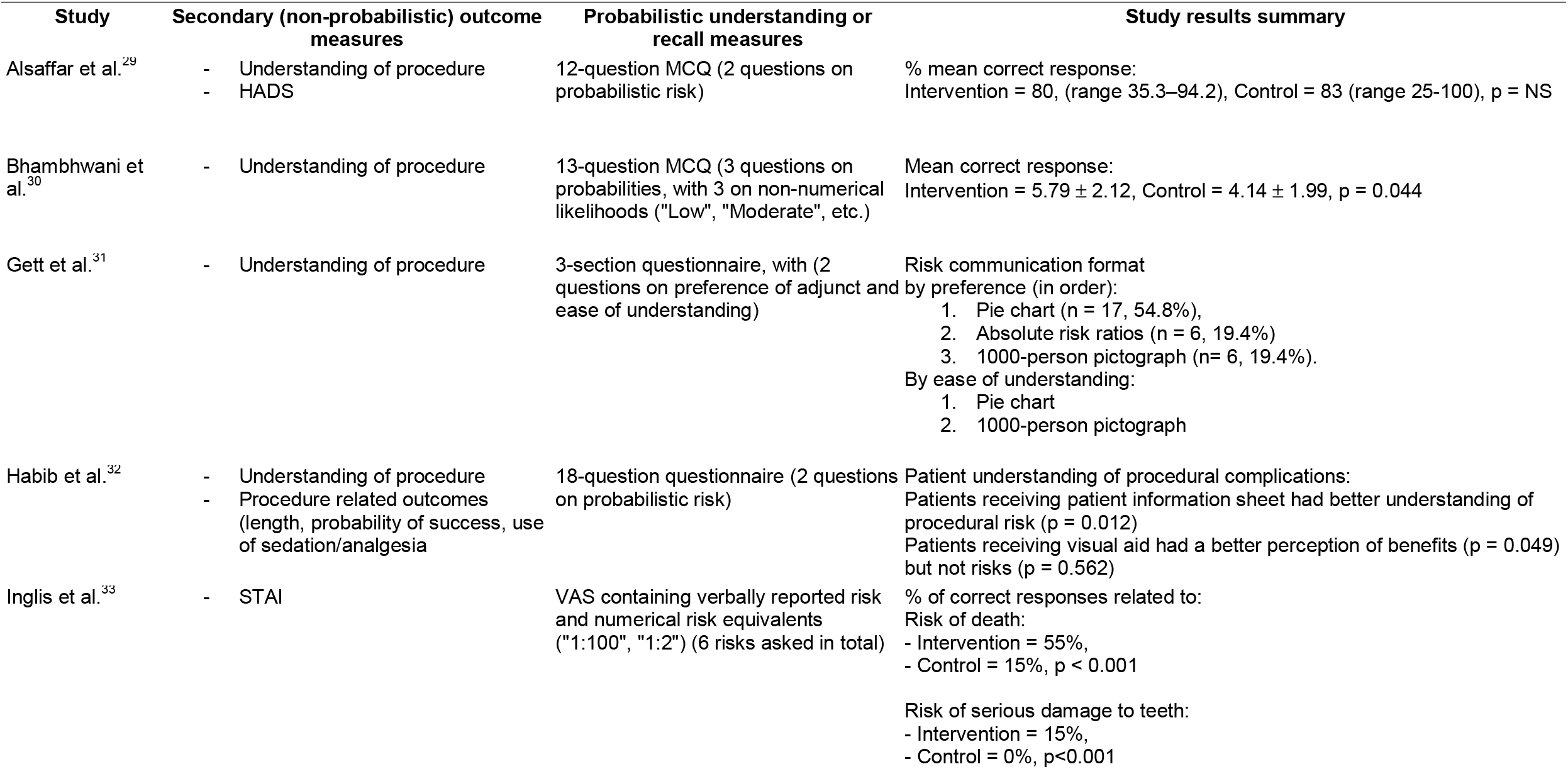
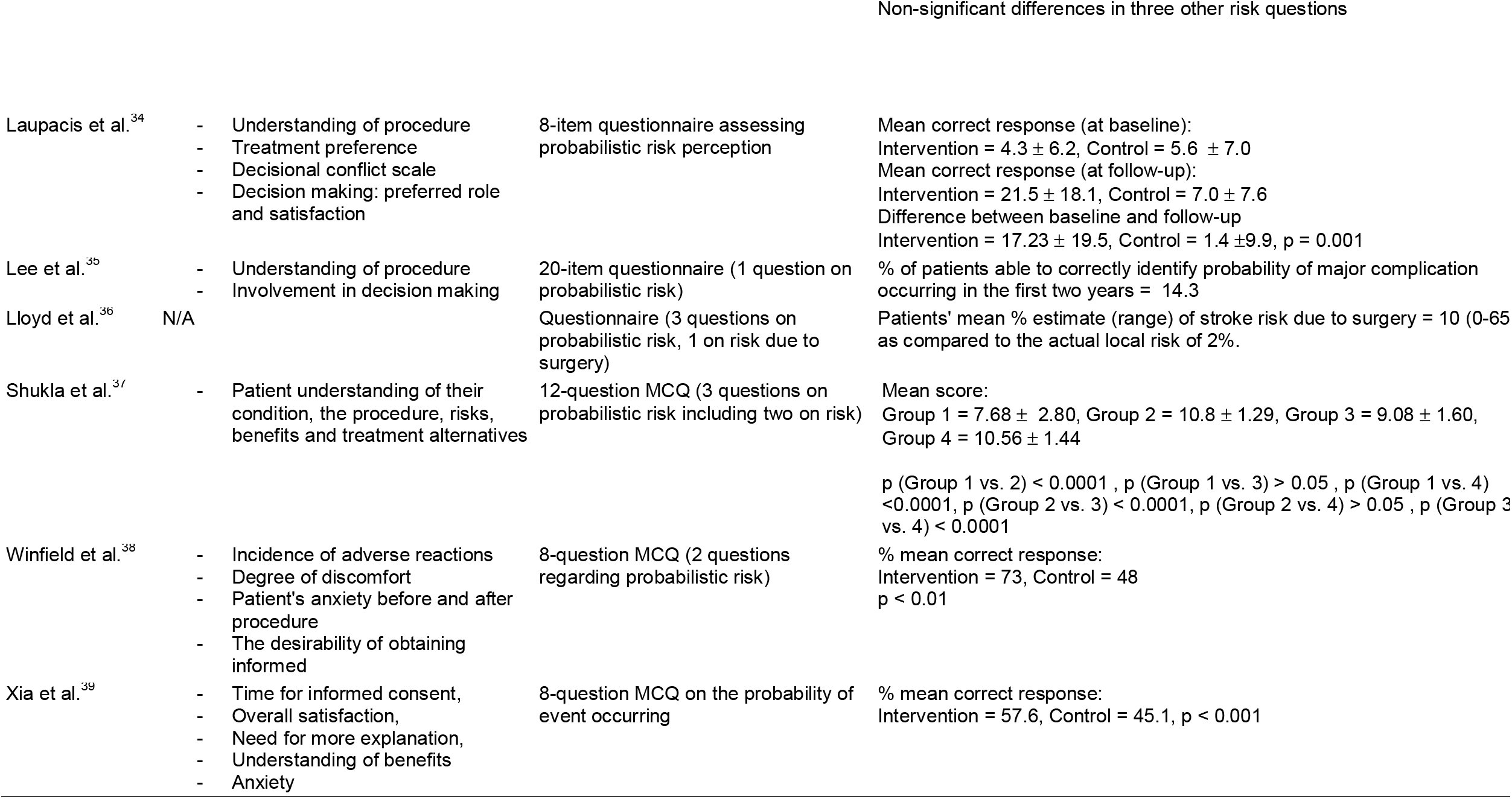
Outcome measures of included studies. HADS = Hospital Anxiety and Depression Scale, STAI = Spielberger State-Trait Anxiety Inventory, MCQ = Multiple choice questionnaire, NS = non-significant, VAS = Visual analogue scale

| Study | Secondary (non-probabilistic) outcome measures | Probabilistic understanding or recall measures | Study results summary |
| --- | --- | --- | --- |
| Alsaffar et al.<sup>29</sup> | - Understanding of procedure<br>- HADS | 12-question MCQ (2 questions on probabilistic risk) | % mean correct response:<br>Intervention = 80, (range 35.3–94.2), Control = 83 (range 25-100), p = NS |
| Bhambhwani et al.<sup>30</sup> | - Understanding of procedure | 13-question MCQ (3 questions on probabilities, with 3 on non-numerical likelihoods ("Low", "Moderate", etc.) | Mean correct response:<br>Intervention = 5.79 ± 2.12, Control = 4.14 ± 1.99, p = 0.044 |
| Gett et al.<sup>31</sup> | - Understanding of procedure | 3-section questionnaire, with (2 questions on preference of adjunct and ease of understanding) | Risk communication format by preference (in order):<br>1. Pie chart (n = 17, 54.8%),<br>2. Absolute risk ratios (n = 6, 19.4%)<br>3. 1000-person pictograph (n= 6, 19.4%).<br>By ease of understanding:<br>1. Pie chart<br>2. 1000-person pictograph |
| Habib et al.<sup>32</sup> | - Understanding of procedure<br>- Procedure related outcomes (length, probability of success, use of sedation/analgesia) | 18-question questionnaire (2 questions on probabilistic risk) | Patient understanding of procedural complications:<br>Patients receiving patient information sheet had better understanding of procedural risk (p = 0.012)<br>Patients receiving visual aid had a better perception of benefits (p = 0.049) but not risks (p = 0.562) |
| Inglis et al.<sup>33</sup> | - STAI | VAS containing verbally reported risk and numerical risk equivalents ("1:100", "1:2") (6 risks asked in total) | % of correct responses related to:<br>Risk of death:<br>- Intervention = 55%,<br>- Control = 15%, p < 0.001<br>Risk of serious damage to teeth:<br>- Intervention = 15%,<br>- Control = 0%, p<0.001 |
| Non-significant differences in three other risk questions |  |  |  |
| Laupacis et al.<sup>34</sup> | - Understanding of procedure<br>- Treatment preference<br>- Decisional conflict scale<br>- Decision making: preferred role and satisfaction | 8-item questionnaire assessing probabilistic risk perception | Mean correct response (at baseline):<br>Intervention = $4.3 \pm 6.2$, Control = $5.6 \pm 7.0$<br>Mean correct response (at follow-up):<br>Intervention = $21.5 \pm 18.1$, Control = $7.0 \pm 7.6$<br>Difference between baseline and follow-up<br>Intervention = $17.23 \pm 19.5$, Control = $1.4 \pm 9.9$, $p = 0.001$ |
| Lee et al.<sup>35</sup> | - Understanding of procedure<br>- Involvement in decision making | 20-item questionnaire (1 question on probabilistic risk) | % of patients able to correctly identify probability of major complication occurring in the first two years = 14.3 |
| Lloyd et al.<sup>36</sup> | N/A | Questionnaire (3 questions on probabilistic risk, 1 on risk due to surgery) | Patients' mean % estimate (range) of stroke risk due to surgery = 10 (0-65) as compared to the actual local risk of 2%. |
| Shukla et al.<sup>37</sup> | - Patient understanding of their condition, the procedure, risks, benefits and treatment alternatives | 12-question MCQ (3 questions on probabilistic risk including two on risk) | Mean score:<br>Group 1 = $7.68 \pm 2.80$, Group 2 = $10.8 \pm 1.29$, Group 3 = $9.08 \pm 1.60$, Group 4 = $10.56 \pm 1.44$<br>$p$ (Group 1 vs. 2) < 0.0001, $p$ (Group 1 vs. 3) > 0.05, $p$ (Group 1 vs. 4) < 0.0001, $p$ (Group 2 vs. 3) < 0.0001, $p$ (Group 2 vs. 4) > 0.05, $p$ (Group 3 vs. 4) < 0.0001 |
| Winfield et al.<sup>38</sup> | - Incidence of adverse reactions<br>- Degree of discomfort<br>- Patient's anxiety before and after procedure<br>- The desirability of obtaining informed | 8-question MCQ (2 questions regarding probabilistic risk) | % mean correct response:<br>Intervention = 73, Control = 48<br>$p < 0.01$ |
| Xia et al.<sup>39</sup> | - Time for informed consent,<br>- Overall satisfaction,<br>- Need for more explanation,<br>- Understanding of benefits<br>- Anxiety | 8-question MCQ on the probability of event occurring | % mean correct response:<br>Intervention = 57.6, Control = 45.1, $p < 0.001$ |

A range of specialties were represented in our findings with only ophthalmology being represented in more than one study^28,29^. In addition to patients undergoing surgery, four non-surgical (but consent requiring) interventional procedures were included: excretory urography, endoscopic retrograde cholangiopancreatography colonoscopy and angioplasty.

The interventions studied could primarily be divided into three types: (i) written information relating to risk information (n = 3, 33.3%); (ii) graphical presentations of risk (n = 1, 11.1%), (iii) Audio/video tools such as audio tapes or DVDs and online videos (n = 11.1%). Another four studies (44.4%) compared multiple interventions or used a blended intervention (e.g. audiotape with written information) Two studies compared multiple interventions: Shukla et al. (written brochures and DVDs) ^29^ and Gett et al.^30^ (written text and visual representations) while Laupacis et al. used a blended intervention using an audiotape and booklet^31^.

The majority of studies assessed the impact of interventions in improving patients’ knowledge of a particular procedure through a multiple-choice questionnaire. As the aim of our review was on interventions to improve probabilistic understanding of risk, study questionnaires were required to have questions pertaining to probability. The number of questions requiring a patient to recall the exact probability of an event varied between studies from 1-8 questions (median = 2.5)^31-33^. Other outcomes of relevance to this review studied included preferred method or risk communication^30^.

### Quality Assessment

Figure 3. presents a summary of the quality assessment which found ‘some concerns’ with all included randomised control trials. This was largely due to issues with outcome reporting, namely a lack of statistical analysis plan and whether the results for questions relating to probabilistic risk could be separated out from overall knowledge, or with the reporting of the randomization process. **Figure 4** presents a summary of the quality assessment for cross-sectional studies, with studies ranging from unsatisfactory to satisfactory. Low scores in selection for Lloyd et al. and Gett et al. for selection were primarily due to the lack of a clear sample-size justification and lack of information on non-respondents. All three studies had issues relating to outcome due to the lack of a validated assessment measure.

Due to the nature of the specific outcomes evaluated in this review, the quality assessment tools used here may not capture the true extent of bias. Not all studies reported the results of probabilistic understanding separately and as a result improvements in knowledge scores could be attributed to questions not pertaining to probabilistic risk.

**Figure 3.**
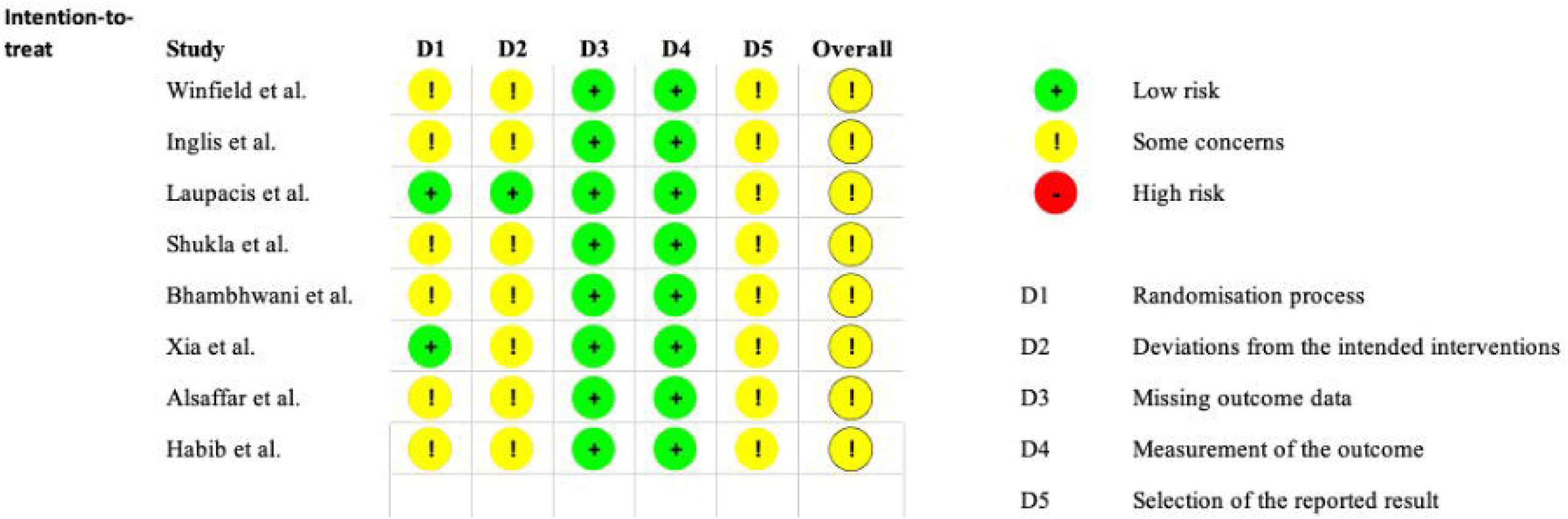
Consensus results of Risk of Bias 2 (RoB2) assessment for randomised control trials

**Figure 4.**
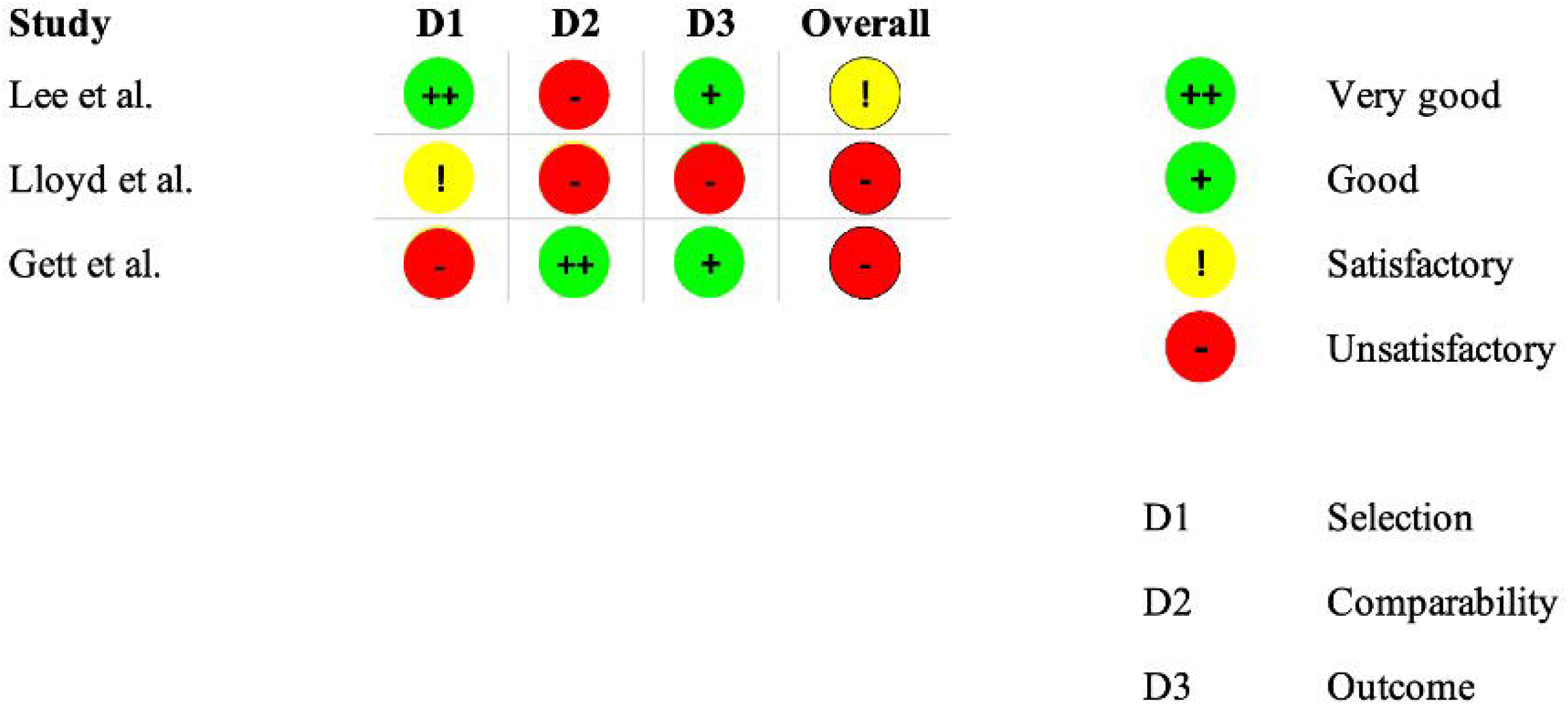
Quality assessment of cross-sectional studies according to modified Newcastle-Ottawa scale. ‘Unsatisfactory’ studies are those scoring below 4, ‘satisfactory’ studies score 5-6, good studies score 7-8 and finally ‘very good’ studies would be scored 9-10

### Findings

#### Recall and perception of probabilistic risk in observational studies

Two non-interventional studies assessed patients’ recall and perception of probabilistic risk. Lee et al. found patients’ knowledge about complications for mastectomy was especially low with only 14.3% being able to accurately recall the correct probability of a major complication, with most patients thinking it was lower than the actual risk ^34^. Lloyd et al. found patients were also inaccurate about the stroke risk associated with carotid surgery with 23% of patients unable to answer the question at all. In this study patients significantly overestimated the risk of stroke, and this was dependent on interval between assessment and surgery date. On the day before the procedure, patients estimated mean stroke risk to be almost 3 times greater than the mean risk estimated a month after their original clinic appointment where surgery was first offered ^35^.

#### Written information

Seven studies utilised written information as a risk communication device with two studies using a blended intervention of an audiotape in addition to written information (discussed in the next section)^31^. Winfield et al., Bhambhwani et al. and Alsaffar et al. compared the use of written information through patient information sheets with standard verbal information^28,36,37^. Winfield et al., and Bhambwani et al. found statistically significant improvements in the intervention as compared to the control group in the overall mean score on the knowledge-based questionnaire^28,36^. Alsaffar et al., in contrast, found no statistically significant differences. In this study, although a greater proportion of participants answered correctly in two of the three questions related to probability, these differences were non-significant^37^.

In Habib et al., patients were assessed on their knowledge of peripheral angioplasty with a subset of patients having received an additional patient information sheet which included information on the procedure, rates of success, associated risks and benefits^33^. Those who had received the patient information sheet were found to have more realistic perceptions as measured by a patient questionnaire of overall risk as well as the individual risks of the procedure^33^.

Shukla et al. compared the use of a patient information sheet at two different reading levels (second and eighth grade) against groups either receiving the standard verbal information or watching a 13-minute DVD. Patients at the second-grade reading level information sheet group scored significantly higher on the knowledge-based questionnaire than both the control group and eighth-grade reading level information sheet group suggesting that presenting the information at a lower reading level or at a more appropriate level for the patient increases ease of understanding.

#### Audio/video

Laupacis et al. and Inglis et al. studied the use of an audio tape (with written information) while Xia et al. studied the use of an educational video compared to ‘routine’ information presented in written or audiotape format ^31,32,38^. Laupacis et al. found that the intervention group overall had significantly improved ability to accurately recall the probability of risks associated with their respective procedures^32^. The impact of an audiotape/written information was more limited in Inglis et al. with the intervention group being able to better recall probabilistic risk of only the rarer rather than common complications ^38^. In both Laupacis et al. and Xia et al., across each individual probability they were asked to recall, the intervention groups had a greater recall of risk incidence as compared to controls. For individual risk, Laupacis et al. noted significant differences in the intervention’s group’s ability to accurately recall the incidence of 6 out of 8 risks, with Xia et al. finding significant differences in probabilistic risk recall for 5 out of 8 risks^31,32^. It should be noted that the improvements in recall in Laupacis et al. were at a mean follow up of 10.0 days showing sustained effects of the risk communication tool used.

Shukla et al. included a patient educational video as an intervention of interest. Patients in the video group had significantly improved overall knowledge scores compared to both the conventional information group and eighth grade reading level brochure group but not the second-grade reading level brochure group. Although no overall statistically significant difference was found between patients in the second-grade reading brochure group and the video group, the prior outperformed the video group in each individual risk related question, highlighting the importance of presenting information at a level of that can be understood by the patients is just as, important as the medium used ^29^.

#### Visual and graphical representation of risks

Gett et al^30^. compared patient preferences and ease of understanding for communicating the risk of perforation during colonoscopy using five different methods of risk communication: (i) absolute risk ratios compared to ‘everyday’ risks (e.g., road traffic accident); (ii) relative risk ratios compared to ‘everyday’ risks; (iii) pie chart; (iv) 1000-person pictograph and (v) a logarithmic scale. The pie chart and 1000-person pictograph were significantly easier to understand than both written forms of risk communication (absolute risk and relative risk) and graphical logarithmic risk scale. The most preferred risk communication format was the pie chart followed by absolute risk ratios as ranked by patients.

In Habib et al, patients undergoing peripheral angioplasty were randomised to a group using a risk assessment chart when answering questions on peripheral angioplasty or without a risk assessment chart. Use of the chart was found to significantly alter patients’ perception of the procedural benefits but not complications.

## DISCUSSION

### Summary

We present a systematic review of interventions aimed at improving probabilistic understanding of risk in patients undergoing interventional procedures. Overall, studies were largely heterogenous in terms of intervention and outcome assessment. Knowledge of probabilistic risk information was found to be poor in observational studies with generally low recall of discussed numerical risks. The majority of RCTs concluded aids in the form of written or audio/visual interventions could improve patients’ ability to recall probabilistic risk. However, except for a few studies which addressed this exclusively, this was based on a limited set of questions. Four studies assessing written information found statistically significant improvements in the understanding of probabilistic risk compared to the standard informed consent proess^28,29,33,36^. Likewise, four studies assessing the use of audio-video tools also found statistically significant improvements compared to the standard information process used in their practice.^29,31,32,38^.

No specific type of medium of decision can be concluded as being the most effective in terms of retention. Indeed, only a single study compared multiple decision aids with no statistically significant differences between a video and written information group^29^. Although, there was limited evidence of patient preference of risk communication tools, one moderately-sized cross-sectional study did find there were preferences for presentations of risk used; with a preference for pie charts over absolute risk ratios and pictographs^30^.

### Study quality

Although these findings show that the use of additional risk communication methods appears to improve the understanding of statistical risk information, our findings should be treated with caution. Outcome measurement between studies was variable, with large differences in the number of questions relating to statistical risk. Furthermore, not all studies analysed the questions relating to statistical risk information separately from the overall knowledge-based questionnaire. As a result, differences in knowledge scores may be attributable to knowledge of other aspects of the procedure other than their probabilistic risk. Despite this limitation, Inglis et al., Laupacis et al. and Xia et al. which all analysed probabilistic risk recall as their sole outcome, all found statistically significant improvements among the intervention groups ^31,38^. Although study size also varied considerably (28 - 205 patients), the two largest trials, Xia et al. and Habib et al. both found significant differences in probabilistic risk recall in patients receiving a risk communication tool^32,33^.

As most studies only assessed numerical understanding of the procedure at a single timepoint, we are unable to conclude the impact of decision aids on long term retention. Furthermore, it is unclear how long the patient is exposed to the risk communication device or whether they may be able to refer to it during assessment. Certainty of evidence was, at least in part, downgraded due to concerns in risk of bias assessment of studies and indirectness of certain studies where answers to questions relating to probabilistic risk was not analysed separately. This led to a final rating of low quality (**Table 3.)**

**Table 3.**
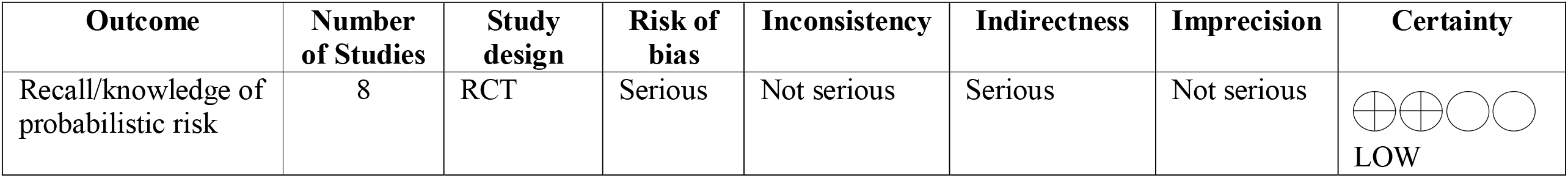
Certainty of evidence for our outcome using GRADE

| Outcome | Number of Studies | Study design | Risk of bias | Inconsistency | Indirectness | Imprecision | Certainty |
| --- | --- | --- | --- | --- | --- | --- | --- |
| Recall/knowledge of probabilistic risk | 8 | RCT | Serious | Not serious | Serious | Not serious | LOW |

### Interpretation and context

In other studies which assess the recall of procedural complications without associated probabilities, the provision of information in written or video form has already been shown to improve the ability of patients to recall information ^15,39,40,41^. By including numerical information, consent aids may go further in helping patients weigh up their decision to undergo a particular procedure. Estimations against a spectrum of common and uncommon ‘real-life’ risks allow patients to compare the risk of the procedure and relative incidence of complications against their individual acceptable risk thresholds.

Since an adequate level of numeracy is required with appropriate heuristics to navigate probabilistic information, surgeons and interventionalists should be aware of their patient’s level of numerical literacy. This was demonstrated by Shukla et al. which found information delivered at a lower reading level led to greater probabilistic recall compared to that at a higher reading level. Patient preferences for their method of risk communication should also be considered rather than adopting the same approach for each patient. While pie charts and bar charts are a generally preferred presentation^30,42^, icon arrays have been one such tool shown to specifically help communicate medical risk to patients with low numeracy^43,44^.

Managing pre-procedural patient expectations is directly relevant to patient outcomes following surgery. Fulfilment of patient expectations has shown some association with improved patient-reported outcomes^45^ while understanding that things may go wrong can help manage anxiety and distress if a complication occurs. Clinicians may not provide the individual probabilities of complications occurring to patients is for fear it may cause unnecessary anxiety with implications for post-operative recovery^46^. Some mismatch between the level at which surgeons and patients deem necessary to disclose has previously been found. Wolf et al. found most surgeons believed a 1% threshold was deemed necessary for risk disclosure despite 69% of patients having a threshold of 0.1-1% ^47,48^. In a study of sinus surgery found 85% of patients wanted to know all the risks involved with the procedure regardless of frequency^49^. Though not one of our initial outcomes, four of the included studies measured patient anxiety as an outcome, with all four finding no significant difference between intervention and control groups^32,36-38^ and two of these demonstrating this using validated anxiety scales^50,51^. This suggests that clinicians can disclose the necessary information regarding complications and their associated probabilistic information to support informed decision making without fear it may cause anxiety in patients.

The extent to which risk communication tools are used in actual practice is unclear. Concerns may include slowing down the consent process if the clinician would need to explain a diagram rather than verbally communicating a risk in a single sentence. Risk communication tools could be implemented at multiple timepoints in the consent process. Providing information prior to consenting patients may guide discussion, while providing information after will allow them to reference information continuously. The ability to recall information about procedures, including complications without associated probabilities, has been shown to decrease over time and may require opportunities for re-discussion ^52-54^. Laupacis et al. was the only study measuring outcomes at two different timepoints (with an average interval of 10.0 days), finding those who had received the intervention had better retention of probabilistic risk at follow-up after their consent for the procedure was first taken^31^. With respect to understanding of a procedure itself, it has been found improvements in knowledge about a procedure were maintained at day of surgery before returning to baseline at 6 weeks post operatively^41^.

Electronic consent (eConsent) forms represents a multimodal method of consent that is currently being trialled in academic research settings^55^ and have been shown to improve the quality of documentation in surgical consent^56^. Risk communication tools such as audio-visual tools providing information of a procedure alongside interactive functionality can be directly embedded within an eConsent form for ease of viewing and has been shown to enhance understanding^55^. As demonstrated by Gett et al., patient preference for how risk is presented varies^30^. Like audio-visual tools, eConsent would be ideally placed to incorporate personalized risk communication formats, allowing patients to readily choose from a range of formats according to their preference and ability.

The need to disclose risks relevant to the individual patient has existed in the USA since Canterbury v. Spence where a patient was not sufficiently warned of the risk of paralysis^57^. It was deemed a risk was material when “a reasonable person, in what the doctors knows or should know to be the patient’s position, would be likely to attach significance to the risk”^57^. A similar ruling was recently passed down in the United Kingdom in Montgomery v Lanarkshire Health Board^58^. Risk communication tools can potentially act as a reminder of complications discussed during the consent process. This could be through specifying the list of risks that need to be included for a particular procedure; allowing some form of standardization with the risks discussed while allowing for flexibility in how they are presented. Ultimately, those responsible for designing such tools will still need to ensure that all material risks are included to reduce the likelihood of litigation.

### Review strengths and limitations

The strengths of our review include a comprehensive search of five databases, the diverse number of interventional specialties included, and the range of decision aid methods represented. However, to ensure a comprehensive search and using our definition of an intervention we have also included studies that are more proximal to medicine and radiology as versus surgery. There was also significant diversity in terms of outcome measurements and reporting. Studies assessed recall for a different number of risks with not every study reporting the results of these questions separately from overall knowledge scores. As a result, we were unable to perform a meta-analysis on the data to quantify the effects of the different tools.

Future research in this field should aim to compare multiple decision aids or tools in a single population or setting through a randomised control trial to determine the decision aids patients respond to the most. Baseline measurements of health literacy and numeracy should also be considered to assess for any differences or preferences with varying levels. Additionally, decision aids can be treated as working synergistically rather than individually to maximize the strengths of individual decision aid tools. For example, the incorporation of visual representations of risk such as icon arrays into patient information sheets or provision of both a video and written information may have greater effectiveness and appeal to a greater section of patients. Outcome measurement is also another potential area of further research. Development of validated tools to measure patient understanding of probabilistic risk of complications in surgery will allow for comparison between study populations and a future meta-analysis to quantify the impact of decision aids and risk communication tools.

### Conclusion

This systematic review found there is evidence that risk communication tools can improve patient understanding of probabilistic risk. However, given the concerns of quality of the studies these findings should be treated cautiously. Moreover, given the number of studies and varied outcome measurement and reporting we are unable to determine the most effective tool and future randomised controlled trials are urgently needed in this regard.

Ultimately, the tool that is most effective is likely to be dependent on the individual patient’s needs and preferences. As with other areas of medicine, healthcare professionals will need to take a personalised approach in ensuring patients’ expectations are managed appropriately.

## Supporting information

Supplementary 1. (Search Strategy)

Supplementary 2. (PRiSMA Abstract checklist)

Supplementary 3. (PRISMA Abstract Checklist)

## Data Availability

All data produced in the present study are available upon reasonable request to the authors

## Acknowledgements

The authors would like to acknowledge the contribution of Kate Brunskill in developing and performing the systematic search used in this study, Noor Adeebah Mohamed Razif for her comments during the editing of this manuscript.

