## Supplementary 1. (Search Strategy) for "Aids to Improve Statistical Risk Communication in Patients Consenting for Surgery and Interventional Procedures: a Systematic Review"

### Date of search

- 13 July 2021

### Sources searched

- Medline - Ovid MEDLINE(R) ALL 1946 to July 12, 2021
- Embase - Embase 1974 to 2021 July 12
- PsycInfo - APA PsycInfo 1806 to July Week 1 2021
- Scopus
- Web of Science - Science Citation Index Expanded (SCI-EXPANDED), Social Sciences Citation Index (SSCI)

### Search syntax

Ovid MEDLINE(R) ALL <1946 to July 12, 2021> (link to search)

| | |
|---|---|
| 1 | ((literac* or understand* or perceiv* or perception* or comprehen* or knowledg* or decide or decision*) adj4 (numer* or probab* or statist* or math* or number* or odds or percent* or chance or data or evidence or certain* or uncertain*)).mp. |
| 2 | ((surgery or surgical or intervent* or procedure* or operat*) adj4 (risk* or adverse or complicat*)).mp. |
| 3 | (patient* or participant* or candidat* or laypeople or layperson or lay person* or lay people or public or individual*).mp. |
| 4 | 1 and 2 and 3 |
| 5 | Comprehension/ |
| 6 | Decision Making/ |
| 7 | consumer health information/ |
| 8 | health literacy/ |
| 9 | exp Informed Consent/ |
| 10 | or/5-9 |
| 11 | mathematics/ |
| 12 | exp statistics as topic/ |
| 13 | exp probability/ |
| 14 | uncertainty/ |
| 15 | exp Evidence-Based Practice/ |
| 16 | or/11-15 |
| 17 | exp Specialties, Surgical/ |
| 18 | exp Surgical Procedures, Operative/ |
| 19 | or/17-18 |
| 20 | exp Postoperative Complications/ |
| 21 | exp risk/ |
| 22 | Long Term Adverse Effects/ |
| 23 | or/20-22 |
| 24 | exp Patients/ |
| 25 | attitude to health/ or health knowledge, attitudes, practice/ or "treatment adherence and compliance"/ or exp "patient acceptance of health care"/ or exp patient satisfaction/ or exp treatment refusal/ |
| 26 | Patient Education as Topic/ |
| 27 | or/24-26 |

| | |
|---|---|
| 28 | 10 and 16 and 19 and 23 and 27 |
| 29 | 4 or 28 |
| 30 | Epidemiologic Studies/ |
| 31 | exp Case-Control Studies/ |
| 32 | exp Cohort Studies/ or Cross-Sectional Studies/ |
| 33 | case control.tw. |
| 34 | ((cohort or epidemiologic$) adj (study or studies)).tw. |
| 35 | cohort analy$.tw. |
| 36 | (follow up adj (study or studies)).tw. |
| 37 | (observational adj (study or studies)).tw. |
| 38 | longitudinal.tw. |
| 39 | retrospective$.tw. |
| 40 | prospective$.tw. |
| 41 | cross sectional.tw. |
| 42 | or/30-41 |
| 43 | Randomized Controlled Trials as Topic/ |
| 44 | randomized controlled trial/ |
| 45 | Random Allocation/ |
| 46 | Double Blind Method/ |
| 47 | Single Blind Method/ |
| 48 | clinical trial/ |
| 49 | clinical trial, phase i.pt. |
| 50 | clinical trial, phase ii.pt. |
| 51 | clinical trial, phase iii.pt. |
| 52 | clinical trial, phase iv.pt. |
| 53 | controlled clinical trial.pt. |
| 54 | randomized controlled trial.pt. |
| 55 | multicenter study.pt. |
| 56 | clinical trial.pt. |
| 57 | exp Clinical Trials as topic/ |
| 58 | (clinical adj trial$).tw. |
| 59 | ((singl$ or doubl$ or trebl$ or tripl$) adj (blind$3 or mask$3)).tw. |
| 60 | Placebos/ |
| 61 | placebo$.tw. |
| 62 | randomly allocated.tw. |
| 63 | (allocated adj2 random$).tw. |
| 64 | or/43-63 |
| 65 | case report.tw. |
| 66 | letter/ |
| 67 | historical article/ |
| 68 | or/65-67 |
| 69 | 64 not 68 |
| 70 | 42 or 69 |
| 71 | 29 and 70 |

Embase <1974 to 2021 July 12> (link to search)

| | |
|---|---|
| 1 | ((literac* or understand* or perceiv* or perception* or comprehen* or knowledg* or decide or decision*) adj4 (numer* or probab* or statist* or |

| | |
|---|---|
| | math* or number* or odds or percent* or chance or data or evidence or certaint* or uncertaint*)).ti,ab. |
| 2 | ((surgery or surgical or intervent* or procedure* or operat*) adj4 (risk* or adverse or complicat*)).ti,ab. |
| 3 | (patient* or participant* or candidat* or laypeople or layperson or lay person* or lay people or public or individual*).ti,ab. |
| 4 | 1 and 2 and 3 |
| 5 | comprehension/ |
| 6 | exp decision making/ |
| 7 | consumer health information/ |
| 8 | health literacy/ |
| 9 | informed consent/ |
| 10 | or/5-9 |
| 11 | mathematics/ |
| 12 | exp statistics/ |
| 13 | exp statistical concepts/ |
| 14 | exp evidence based practice/ |
| 15 | or/11-14 |
| 16 | exp surgery/ |
| 17 | exp surgical technique/ |
| 18 | or/16-17 |
| 19 | exp postoperative complication/ |
| 20 | exp risk/ |
| 21 | 19 or 20 |
| 22 | exp patient/ |
| 23 | exp patient attitude/ |
| 24 | patient education/ |
| 25 | or/22-24 |
| 26 | 10 and 15 and 18 and 21 and 25 |
| 27 | 4 or 26 |
| 28 | clinical study/ |
| 29 | case control study/ |
| 30 | family study/ |
| 31 | longitudinal study/ |
| 32 | retrospective study/ |
| 33 | prospective study/ |
| 34 | cohort analysis/ |
| 35 | (cohort adj (study or studies)).ti,ab. |
| 36 | (case control adj (study or studies)).ti,ab. |
| 37 | (follow up adj (study or studies)).ti,ab. |
| 38 | (observational adj (study or studies)).ti,ab. |
| 39 | (epidemiologic* adj (study or studies)).ti,ab. |
| 40 | retrospective$.ti,ab. |
| 41 | prospective$.ti,ab. |
| 42 | (cross sectional adj (study or studies)).ti,ab. |
| 43 | or/28-42 |
| 44 | Clinical Trial/ |
| 45 | Randomized Controlled Trial/ |
| 46 | controlled clinical trial/ |
| 47 | multicenter study/ |

| | |
|---|---|
| 48 | Phase 3 clinical trial/ |
| 49 | Phase 4 clinical trial/ |
| 50 | exp randomization/ |
| 51 | single blind procedure/ |
| 52 | double blind procedure/ |
| 53 | crossover procedure/ |
| 54 | placebo/ |
| 55 | randomi?ed controlled trial$.ti,ab. |
| 56 | (random$ adj2 allocat$).ti,ab. |
| 57 | single blind$.ti,ab. |
| 58 | double blind$.ti,ab. |
| 59 | ((treble or triple) adj blind$).ti,ab. |
| 60 | placebo$.ti,ab. |
| 61 | prospective study/ |
| 62 | or/44-61 |
| 63 | case study/ |
| 64 | case report.ti,ab. |
| 65 | abstract report/ or letter/ |
| 66 | conference proceeding.pt. |
| 67 | conference abstract.pt. |
| 68 | editorial.pt. |
| 69 | letter.pt. |
| 70 | note.pt. |
| 71 | or/63-70 |
| 72 | 62 not 71 |
| 73 | 43 or 72 |
| 74 | 27 and 73 |

APA PsycInfo <1806 to July Week 1 2021> (link to search)

| | |
|---|---|
| 1 | ((literac* or understand* or perceiv* or perception* or comprehen* or knowledg* or decide or decision*) adj4 (numer* or probab* or statist* or math* or number* or odds or percent* or chance or data or evidence or certain* or uncertain*)).mp. |
| 2 | ((surgery or surgical or intervent* or procedure* or operat*) adj4 (risk* or adverse or complicat*)).mp. |
| 3 | (patient* or participant* or candidat* or laypeople or layperson or lay person* or lay people or public or individual*).mp. |
| 4 | 1 and 2 and 3 |
| 5 | exp Comprehension/ |
| 6 | exp Decision Making/ |
| 7 | health literacy/ |
| 8 | exp Informed Consent/ |
| 9 | or/5-8 |
| 10 | mathematics/ |
| 11 | exp statistics/ or exp analysis/ |
| 12 | exp probability/ or exp judgment/ |
| 13 | uncertainty/ |
| 14 | exp evidence based practice/ |

| | |
|---|---|
| 15 | or/10-14 |
| 16 | exp surgery/ |
| 17 | Postsurgical Complications/ |
| 18 | risk assessment/ |
| 19 | 17 or 18 |
| 20 | exp Patients/ |
| 21 | exp client attitudes/ |
| 22 | Client Education/ |
| 23 | or/20-22 |
| 24 | 9 and 15 and 16 and 19 and 23 |
| 25 | 4 or 24 |

Scopus (link to search)

TITLE-ABS-KEY
( ( ( literac* OR understand* OR perceiv* OR perception* OR comprehen* OR knowledg* OR d ecide OR decision* ) W/3 ( numer* OR probab* OR statist* OR math* OR number* OR odds OR percent* OR chance OR data OR evidence OR certain* OR uncertain* ) ) AND ( ( surgery O R surgical OR intervent* OR procedure* OR operat* ) W/3 ( risk* OR adverse OR complicat* ) ) AND ( patient* OR participant* OR candidat* OR laypeople OR layperson OR {lay person*} OR {lay people} OR public OR individual* ) )

Web of Science - Science Citation Index Expanded (SCI-EXPANDED), Social Sciences Citation Index (SSCI) (link to search)

(Topic) ((literac* or understand* or perceiv* or perception* or comprehen* or knowledg* or decide or decision*) NEAR/3 (numer* or probab* or statist* or math* or number* or odds or percent* or chance or data or evidence or certain* or uncertain*)) AND ((surgery or surgical or intervent* or procedure* or operat*) NEAR/3 (risk* or adverse or complicat*)) AND (patient* or participant* or candidat* or laypeople or layperson or lay person* or lay people or public or individual*)
